# Estimating the Changing Infection Rate of COVID-19 Using Bayesian Models of Mobility

**DOI:** 10.1101/2020.08.06.20169664

**Authors:** Luyang Liu, Sharad Vikram, Junpeng Lao, Xue Ben, Alexander D’Amour, Shawn O’Banion, Mark Sandler, Rif A. Saurous, Matthew D. Hoffman

## Abstract

In order to prepare for and control the continued spread of the COVID-19 pandemic while minimizing its economic impact, the world needs to be able to estimate and predict COVID-19’s spread. Unfortunately, we cannot directly observe the prevalence or growth rate of COVID-19; these must be inferred using some kind of model. We propose a hierarchical Bayesian extension to the classic susceptible-exposed-infected-removed (SEIR) compartmental model that adds compartments to account for isolation and death and allows the infection rate to vary as a function of both mobility data collected from mobile phones and a latent time-varying factor that accounts for changes in behavior not captured by mobility data. Since confirmed-case data is unreliable, we infer the model’s parameters conditioned on deaths data. We replace the exponential-waiting-time assumption of classic compartmental models with Erlang distributions, which allows for a more realistic model of the long lag between exposure and death. The mobility data gives us a leading indicator that can quickly detect changes in the pandemic’s local growth rate and forecast changes in death rates weeks ahead of time. This is an analysis of observational data, so any causal interpretations of the model’s inferences should be treated as suggestive at best; nonetheless, the model’s inferred relationship between different kinds of trips and the infection rate do suggest some possible hypotheses about what kinds of activities might contribute most to COVID-19’s spread.

## 1 Introduction

In response to the coronavirus pandemic of 2020, countries around the world instituted non-pharmaceutical interventions in an attempt to slow the spread of the disease. These interventions included such measures as social distancing, mandatory wearing of masks, and shutting down non-essential businesses and services. Understanding the impact of these measures on the spread of the disease is critical to informing decisions and designing interventions, but due to the delay between infection and obtaining test results, it can be weeks before the effect of such an intervention can be seen in case counts and death counts [1]. Ideally, policymakers would have a current estimate (a “nowcast”) showing how the growth rate of the disease is responding to various interventions and other factors, as well as a forecast that quickly adapts to changing conditions and generates “what-if” estimates of how changes in behavior might affect disease spread. Unfortunately, case and death counts are lagging indicators; a new infection may take weeks to lead to a confirmed case and/or death. To draw inferences earlier, we need more-responsive correlates of infection rates.

As part of a wider effort to understand the impact of the non-pharmaceutical interventions, many institutions such as the New York Times [2], Google [3], and Apple [4] have released data about mobility trends. This aggregated mobility information measures movement trends at various levels of granularity (county, state), and across different mobility categories (going to the grocery or work, etc.). Such aggregated mobility data lacks individual-level detail, and may not be a representative sample of the broader population’s behavior, but it may still hold useful signals for understanding the *current* rate of spread of coronavirus. Given our understanding of how coronavirus spreads, we expect that more movement in society results in more infections. We thus hope the mobility trends correlate with the unobserved infection rate of COVID-19. The breakdown of overall mobility into categories based on destination-type or distance also provides an opportunity to gain intuitions about what types of trips most spread the disease (although these intuitions should be treated as hypotheses needing further validation, since the data are observational).

With this available mobility data, the next step is building models that can use it to make reliable predictions. One approach would be to incorporate a mobility signal into a discriminative predictive model. Although it is more difficult to incorporate assumptions and prior knowledge into a discriminative model, the results from [5] indicate that that the correlation between mobility and infection rate is strong enough to make good forecasts. On the other hand, compartmental models [e.g., 1, 6, 7] assume a flexible, causal story for the spread of a disease and can also incorporate mobility data as a covariate for predicting the time-varying infection rate of a disease.

In this paper, we explore the use of mobility data in compartmental models and find that:

- Mobility data can be used to forecast for the spread of the disease.
- Mobility data is a promising low-lag control signal that can be used to infer the invisible spread of COVID-19 and assist policy makers.
- Incorporating mobility data into compartmental models suggests that some types of trips influence the infection rate more than others.

## 2 Background

### 2.1 Compartmental models for epidemiology

Compartmental models for epidemiology partition a population into “compartments” depending on which stage of a disease’s life cycle a member is in. A simple model is the Susceptible-Infectious-Recovered, or SIR model [8], which partitions a population into “Susceptible”, the people who are not yet infected, “Infectious”, the people who have the disease and are actively spreading it to others, and “Recovered”, the people who are no longer infected with the disease, often including both those who successfully recover and those who die from the disease. In practice, we may want to include an “Exposed” compartment, for individuals who have contracted the disease but are not yet infectious, resulting in an SEIR model. An SEIR model assumes an initial state (i.e. some initial numbers (*S*_0_, *E*_0_, *I_0_, R_0_)* and a population size 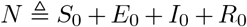 and a set of parameters that govern a simulation that evolves this initial state over time. The parameters include *β*, the infection rate, *α*, the incubation rate, and *γ*, the recovery rate. The evolution of the state over time is prescribed by a set of differential equations: 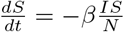; 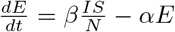; 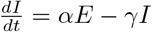; 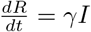.

When using a compartmental model, we are often interested in obtaining the values of each of the compartments at a set of input times. A compartmental model defines a continuous-time process and it is conventional to choose the unit of time to be days. Thus, integrating the equations to obtain the values of each compartment at a set of discrete times *τ* = {0, 1, 2, …, *T*} corresponds to obtaining the values at the beginning of each day for *T* days moving forward from the initial state. We focus on these daily values that are the output of integrating the compartmental model equations; we define “simulating” a compartmental model as the process of obtaining these values via numerical methods like Euler or Runge-Kutta integration.

Formally, we assume a compartmental model is a series of differential equations associated with a set of simulation parameters *θ*. Besides the parameters that govern the differential equations, we also include the initial state s in its parameters. For an SIR model we have *θ* = (*β*, *γ*, *s*); the SEIR model adds an incubation-rate parameter α. The simulation function *M*_SIR_(*θ*) or *M*_SEIR_(*θ*) outputs the values of each compartment at the beginning of a set of days *τ* = {0, 1, …, *T*}. For example, M_SEIR_(*θ*) = (*S*_0:_*_T_, E*_0:_*_T_, I*_0:_*_T_, R*_0:_*_T_*).

### 2.2 Fitting compartmental models

Compartmental models provide a convenient, mechanistic formulation for how a disease spreads in a population. However, most often though we don’t know the parameters of the model beforehand, but we do have some data that can provide a learning signal to fit the parameters, One such signal is the daily number of new cases of a disease, which can be predicted by a compartmental model as the change in *I* + *R* between each day. Formally, assume we observe a time-series of daily case counts *c*_1_*_:T_*. From a set of simulation parameters *θ*, we obtain *I*_0:_*_T_,R*_0:_*_T_* from *M(θ)*, the total number of infected and recovered on each day. We can compute the predicted case counts *ĉ*_1:_*_T_* = (*I* + *R*)_1:_*_T_* − (*I* + *R*)_1:_*_T_*_−1_, and compute a loss *l(c*_1_*_:T_, ĉ*_1_*_:T_*) (e.g. mean-squared error). We can then optimize the parameters with respect to this loss with an algorithm like gradient descent.

Case count data is readily available for COVID-19 but is problematic for a few reasons. First, many (perhaps most) people who are infected are not tested; many mild cases are not tested either because individuals do not know that they are infected or because they have sufficiently mild cases that they either cannot or do not feel the need to get tested. Testing availability and policies are inconsistent across different regions, so the amount of underreporting is presumably also inconsistent across different datasets. Furthermore, demographic differences introduce selection bias: if patients with severe cases are more likely to get tested, then regions with a higher fraction of severe cases (e.g., because they have an older population) will have less underreporting. Second, there can be a significant delay between when tests are administered and when they show up in case counts; this delay is not consistently reported. Finally, there will be false positives and false negatives, and these error rates are also likely to vary by region since training is needed to get a good sample for polymerase chain reaction (PCR) tests.

Death counts are more reliable than case counts and should also correlate with infection rate of the disease, albeit with a larger lag than case counts. We expect there to be a smaller underreporting factor for deaths because it is more unlikely that a death from COVID-19 goes unnoticed than an asymptomatic case. While there will still be false positives and negatives in the data due to misdiagnosed and misreported cases, we expect the death counts to be less noisy and biased than the case counts. Unfortunately, using death counts as a learning signal comes with a significant challenge: time lag. With COVID-19, it may take on the order of 2 to 3 weeks until death after infection, meaning any uptick in infections will not be visible in the death counts for 2 to 3 weeks [7, 9]. This introduces a modeling challenge, where a compartmental model that models the recovery period incorrectly will likely learn incorrect values for the other parameters in order to compensate for the incorrect delay. Furthermore, compartmental models do not usually directly simulate deaths in a population but rather the sum of deaths and proper recoveries. Observing deaths requires introducing a new parameter into the model, *ω*, the infection-fatality-ratio (IFR) in order to convert “recoveries” into the subset that die (and whose deaths are reported as due to COVID-19).

### 2.3 Bayesian models for epidemiology

Several studies have aimed to identify values for the parameters of COVID-19’s behavioral dynamics, such as the lengths of the incubation and recovery periods [9]. Even then, it is unlikely that we can identify a single value for each of these parameters given the differences in methodology and assumptions between studies. However, given the surrounding literature, it is possible to construct a distribution over possible parameters that reflects the uncertainty that comes from both a lack of consensus and an inherently noisy process. A strong prior distribution over parameters also helps with identifiability issues, as adding more parameters to an SIR model often results in nonidentiability.

Bayesian modeling is a flexible framework for incorporating these prior assumptions and capturing the resulting predictive uncertainty. A Bayesian compartmental model proceeds by putting prior distributions over the parameters and initial state of a compartmental model. These priors are opportunities to incorporate external evidence from studies, like observed lengths of incubation and recovery periods. The model assumes that the observed data are sampled with some measurement noise from the output of the simulation given the parameters. Inference about the model parameters proceeds by applying Bayes’s rule.

We define a generative process for an observed death count time series *d*_1:_*_T_*, where *θ* are the parameters of a compartmental model. We define additional parameters *ϕ* = (*r*, *ω*), where *r* is the shape parameter for a negative binomial likelihood, and *ω* the infection-fatality-ratio (IFR).

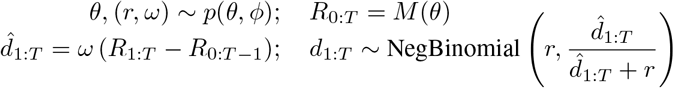

In this generative process, we simulate a compartmental model forward and compute its predicted daily deaths, which is the used as the mean of a noisy observation process. The distributions of interest are the posterior distribution over parameters *p*(*θ*, *ϕ*|*d*_1:_*_T_*) and the posterior predictive distribution *p*(*d*\*|*d*_1:_*_T_*) = *∫ p*(*d*\*|*θ*, *ϕ*)*p*(*θ*, *ϕ*|*d*_1:_*_T_*)*dθdϕ*. The former is useful for exploratory data analysis and provides uncertainty estimates over the parameters in the model and the latter is the distribution needed for forecasting and nowcasting. Unfortunately, we cannot analytically compute these distributions so it is common to use methods that approximate the posterior and predictive distributions like sequential Monte Carlo (SMC) and Markov Chain Monte Carlo (MCMC).

## 3 Extensions to SEIR for COVID-19 modeling

### 3.1 Quarantine compartment and Erlang waiting times

Estimates of the recovery period for COVID-19 are around 15-20 days and we observe this in the lag between observed infections and recoveries. In the basic SEIR model, everyone in the *I* category is infectious at a constant rate for the entire duration of that recovery period. But in reality, it is unlikely that people remain equally infectious during the entirety of the recovery period due to isolation at home and in hospitals. To better model this drop in infectiousness, we add a “*Q*” compartment to the model that contains “isolated” or “quarantined” individuals. Isolated/quarantined individuals behave like infectious ones in that they contribute to the flow from Susceptible to Exposed, but at a fraction of the rate. The new parameter governing the rate of flow between *I* and *Q* we call the “isolation rate” *η* and the fraction governing the reduction in infection rate we call the “quarantine reduction” q. We call this an SEIQR model (visualized in Figure 1) which has the following dynamics: 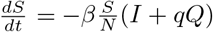; 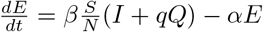; 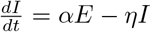; 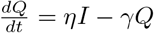; 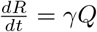.

**Figure 1:**
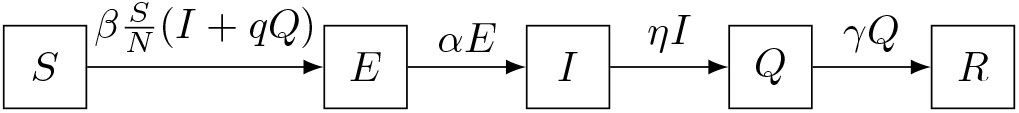
A diagram representing the flow of individuals through an SEIQR models with rates on arrows.

Compartmental models implicitly assume that individuals spend an amount of time in each compartment that is exponentially distributed with that compartment’s rate parameter. An exponential distribution will correctly model an average waiting time, but the overall shape of the waiting time distribution may completely differ from the observed distribution and will put mass on small waiting times. This can be problematic if say we know that a disease often at least 3 days to recover from. A commonly used remedy for the exponential time waiting assumption inherent to SIR and SEIR dynamics is to expand each compartment into several subcompartments and modify the compartment’s rate parameter to match the original’s expected value. A new hyperparameter of our SEIQR model is thus the number of subcompartments for each of the compartments.

### 3.2 Incorporating mobility data

In conventional compartmental modeling, the infection rate parameter *β* is time-invariant, freely parameterized and directly learned from data. This is a strong assumption about how a disease might spread; for example, we might expect that the *true* number of infections on a given day is noisy and that there are likely strong day-of-week effects that come from commuting and varying travel patterns on weekends. (The smoothing effect of the lag between infection and death would make such day-of-week effects invisible in daily death counts.) Furthermore, with COVID-19, we hope that resultant lockdowns cause a drop in the infection rate thereby slowing the spread of the disease. Thus, for modeling COVID-19, we’d like a compartmental model with a time-varying infection rate that captures any drop resulting from non-pharmaceutical interventions.

Mobility data measures the relative change in how many trips are taken daily relative to prelockdown behavior. Each datapoint in mobility data is a *K*-dimensional time-series *m*_0:*T*_; each individual series tracks mobility trends (the higher the values the more trips are being taken, and vice versa). Mobility is further aggregated at the county level, so we have a *K*-dimensional time-series for each of *C* counties 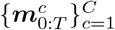.

To model infection rate using mobility data, we define a generalized linear model (GLM) that predicts infection rate. We use a noncentered hierarchical model over GLM weights where we independently sample city-specific base coefficients 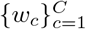. We then sample a normally-distributed *K*-dimensional vector of feature-specific coefficient adjustments ***f*** and sample a base infection rate *b*. We finally compute a county-specific infection rate 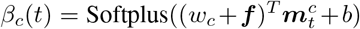. The city coefficients *w_c_* model the differences in how mobility affects infections between different counties and the feature coefficients ***f*** model the infection patterns of different types of mobility.

Mobility is a potential predictor of infection rate because it correlates with the quantity lockdowns are designed to lower- infections coming human-to-human interaction. Some trips may have more human-to-human interactions than other, but in aggregate we expect the numbers of total interactions (and thereby infections) to drop if there are less total trips. Mobility data can be collected and processed relatively quickly and therefore offers the potential to become a signal for forecasting and nowcasting. However, mobility data does not capture all aspects of how disease spreads.

### 3.3 Adding a changepoint

The second modification is incorporating a *changepoint* to model a broader set of latent factors that influence the infection rate. For example, mobility data does not reflect the change in infection rate due to changes in human behavior, such as the widespread wearing of masks and maintaining social distance. These effects are harder to measure but are still relevant to predicting a time-varying infection rate; a changepoint, or a function of time that captures a drop in infection rate, is a convenient means of lumping together all unknown latent factors into a parameterized time-varying infection rate.

We define a changepoint to be a point in time at which there is a monotonic drop in infection rate, modeled as a negative sigmoid. We parameterize it with three parameters: *κ*, or time at which the change happens, *ρ*, a number between 0 and 1 determining the amplitude of the sigmoid, and *ν*, the slope of the sigmoid that dictates the rate of the change. We thus have the parameterized function 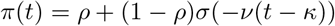 where *σ* is the sigmoid function.

Unlike mobility data, a changepoint cannot model complex changes in infection rate but it can, however, capture factors like differences in human behavior that is not reflected in mobility data. By itself, a changepoint can model the flattening of the death count curves we observe in COVID-19 data (i.e. *β*(*t*) = *π*(*t*)), but a changepoint is not useful for forecasting and nowcasting because we can only infer a changepoint has happened after observing the case or death counts weeks after. Thus, we desire a hybrid model that can leverage the strengths of both approaches to time-varying mobility rate. When modeling multiple counties, we model county-specific changepoints and merge the changepoint with the mobility model by multiplying it by the output of the mobility GLM, i.e. 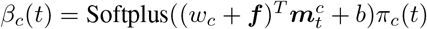.

## 4 Related work

[7] proposed one of the first Bayesian SIR models of COVID-19 based on observed death counts. Rather than treat death as a compartment, they convolve the new-infection curve in a simple SIR-with-observed-changepoint model with a time-to-death distribution. [10] develop a Bayesian semi-mechanistic (rather than compartmental) model linking Google mobility data and a latent time-varying factor to a time-varying reproductive number *R_t_*, which is combined with a latent IFR and time-to-death distribution to model observed deaths. [11] develop a changepoint model similar to ours, informed by lockdown dates rather than mobility data. [12] strongly argue for the importance of relaxing the exponential-waiting-time assumption in compartmental models; see also [13, 14].

Concurrently with this work, [15] proposed a very similar Bayesian compartmental model regressing from Apple mobility data to a time-varying infection rate *β*(*t*). The most salient difference is that we consider a latent changepoint to account for changes in behavior that are not captured by mobility data. We find that this factor is important for making accurate long-term forecasts, although it makes relatively little difference over the 8-day forecast window they consider.

## 5 Experiment setup

### Datasets

We use unprocessed COVID-19 case and death count numbers from the New York Times [2], which produces daily reports of new infections and deaths at both state and county level. For mobility, we use the Community Mobility Reports published by Google [3] in our evaluation of Bayesian compartmental models. Google’s Community Mobility Datasets are created with aggregated, anonymized sets of data from users who have turned on the Location History setting, which is off by default [16]. No personally identifiable information, like an individual’s location, contacts or movement, is made available at any point. Trends are aggregated to the county level (including Washington, DC and independent cities that are not otherwise included in county boundaries), and available daily from February 16th through May 21th, 2020. The Google dataset also breaks down mobility trends into several categories. We consider two usages of the mobility data: (1) using each category as a feature (except for `residential`) and (2) aggregating the categories into a single overall mobility feature.

We normalize the dataset to have 0 represent “normal” mobility and deviations represent positive and negative relative changes in mobility (e.g. a value of 0.3 represents a 30% increase in mobility). Figure 2 shows the daily mobility changes in New York City for each category in Google’s Community Mobility Reports, where “overall” represents the aggregated category. In New York City, we observe a dramatic decrease in many of the mobility categories and in overall mobility after the official non-pharmaceutical intervention occurred in late March. Since mid-April, the number of park visits in New York City has also increased, while the number of visits in other categories is relatively small.

**Figure 2:**
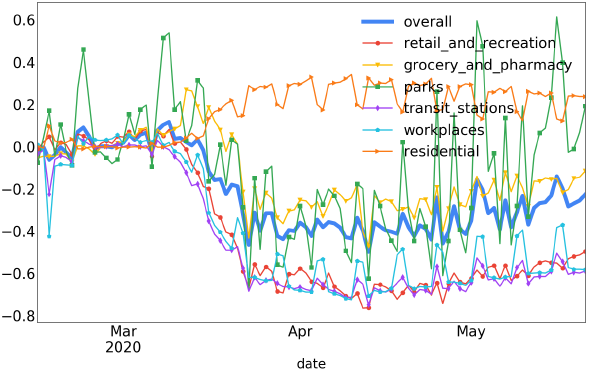
Mobility trends for New York City. There are strong day-of-week effects in the data indicated by the small weekly spikes.

**Figure 3:**
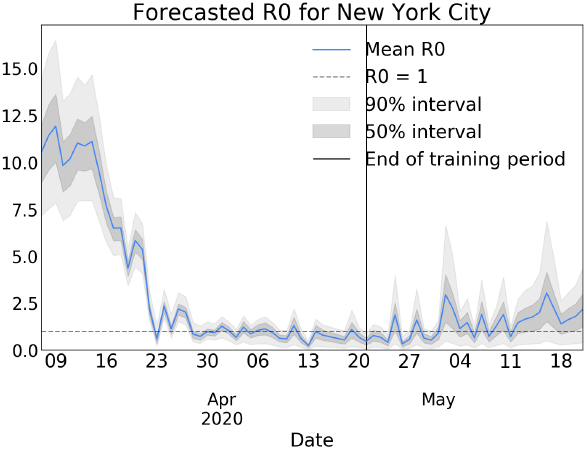
The forecasted value of *R_t_* for New York City according to a CMSEIQR model.

### Model priors and hyperparameters

The hyperparameters of the model are the number of sub-compartments in the Erlang-based models, for which we chose 2 subcompartments for each of the *E*, *I* and *Q* compartments. The priors of the various model parameters are defined in Table A.2. Priors were chosen to match estimates found in [9], which aggregates parameter estimates from relevant literature. The compartmental model parameters (except for infection rate) are shared across counties, and the rest of the parameters in the model are county-specific, with the exception of ***f***, which is mobility-feature specific.

### Numerical integration

We simulate compartmental dynamics with an Euler-discretization of the differential equations with a step size of 1. Despite such a large step size, we found the dynamics to be stable with reasonable parameter settings. However, with extreme values for parameters, we found that the dynamics become unstable and numerical issues arise. We address this by constraining certain parameter values in the prior distribution to avoid extreme values. We also find no significant performance improvement with a lower step size.

### Bayesian inference

We use an adaptive sequential Monte Carlo (SMC) method for Bayesian inference. We utilize the algorithm from [17] which adaptively constructs a series of annealed distributions that interpolates between the prior over latent variables and joint distribution over latent variables. We then initialize a set of particles that are iteratively updated with a Hamiltonian MonteCarlo (HMC) kernel to be representative of each of the annealed distributions [18, 19]. In practice, we map parameters into an unconstrained space and modify the prior to take into account the volume difference resulting from change-of-variable, and we execute SMC in this unconstrained space. We run 5 chains in parallel with 1000 particles each, collecting a total of 5000 samples from the posterior distribution. Inference is implemented using TensorFlow Probability [20, 21] and were executed on a GPU where training a single model took around 20 minutes.

## 6 Results

We focus on the period starting on 3/7/2020 extending to 5/21/2020. We fit various versions of the models described above to the aforementioned mobility trend and ground-truth death counts, and evaluate their ability to forecast, nowcast, and offer insights into how mobility influences COVID-19’s infection rate. To evaluate the extensions proposed in the paper, we we consider a baseline SEIQR model (time-invariant infection rate), a changepoint SEIQR (CSEIQR), an SEIQR model with a infection rate varying as a function of mobility data from both multiple and an aggregated category (MSEIQR[Multi] and MSEIQR[Overall]), and a model that combines both multiple category mobility-based regression and a changepoint (CMSEIQR).

### 6.1 COVID-19 forecasting

To evaluate the overall effectiveness of modeling COVID-19 using the proposed models, we construct a COVID-19 forecasting task on the 25 most populous US counties (treating the five boroughs of New York City as one county). In three variants of this task, we choose a date (4/22/20) and fit a model to the data up to this date. We compute the predictive log-likelihood for the next 7, 14, and 28 days worth of data, using mobility data from the held-out period when forecasting^2^. To measure the quality of each model’s forecasts, we report the average daily marginal log-likelihood numbers for the held-out data, which we estimate by taking the log-mean-exp of of the log-likelihoods for every sample, and averaging across days. This measures how well on average the model can predict the number of deaths on a randomly chosen day in the future.

#### Comparing mobility datasets

We first evaluate whether having access to more fine-grained information about mobility trends is valuable for forecasting. To accomplish this, we examine held-out log-likelihood of MSEIQR[Multi/Overall] models in Table B.4a and Table B.5. We find that the MSEIQR[Multi] consistently achieves a higher average log-likelihood than the MSEIQR[Overall] mobility model in all forecast windows. Investigating further, it appears that the overall mobility- trained model “overfits” to the slow increase in mobility we observe towards the end of the forecast period and predicts upticks in deaths where the data continues to trend downwards. This problem is reduced in the multiple-category model; we thus hypothesize that there are certain types of mobility that better correlate with infection rate and aggregating over different mobility types adds noise to these signals. For example, Figure B.3 shows that despite high uncertainty changes in `grocery_and_pharmacy` are a stronger signal of changes in infection rate than, say, `transit_stations`—this observed association does *not* imply that interventions targeting grocery stores and pharmacies will reduce COVID-19 spread more than closing subways, but it does suggest that these relative changes should not be weighted equally when trying to *estimate* its spread.

#### Comparing model variants

We report held-out log-likelihood numbers for all model variants in subsection B.2. We observe that all mobility models tend to latch onto mobility upticks in in late April and May. MSEIQR[Multi/Overall] models were trained without these upticks and can only explain infection rate in terms of mobility, so subsequently we see a forecasted uptick in deaths for those models. On the other hand, the changepoint model is often correct but every so often, misidentifies the drop in the infection rate and makes a blatantly wrong forecast (see Figure 4). The CMSEIQR model tends to forecast somewhere in between, predicting a decay in death counts but with a slight uptick towards the end of the forecast. However, it has a much wider interval due to the inclusion of the changepoint, which mitigates the effect of mobility towards the end of the forecast period. We visualize the rest of forecasts in subsection C.1.

**Figure 4:**
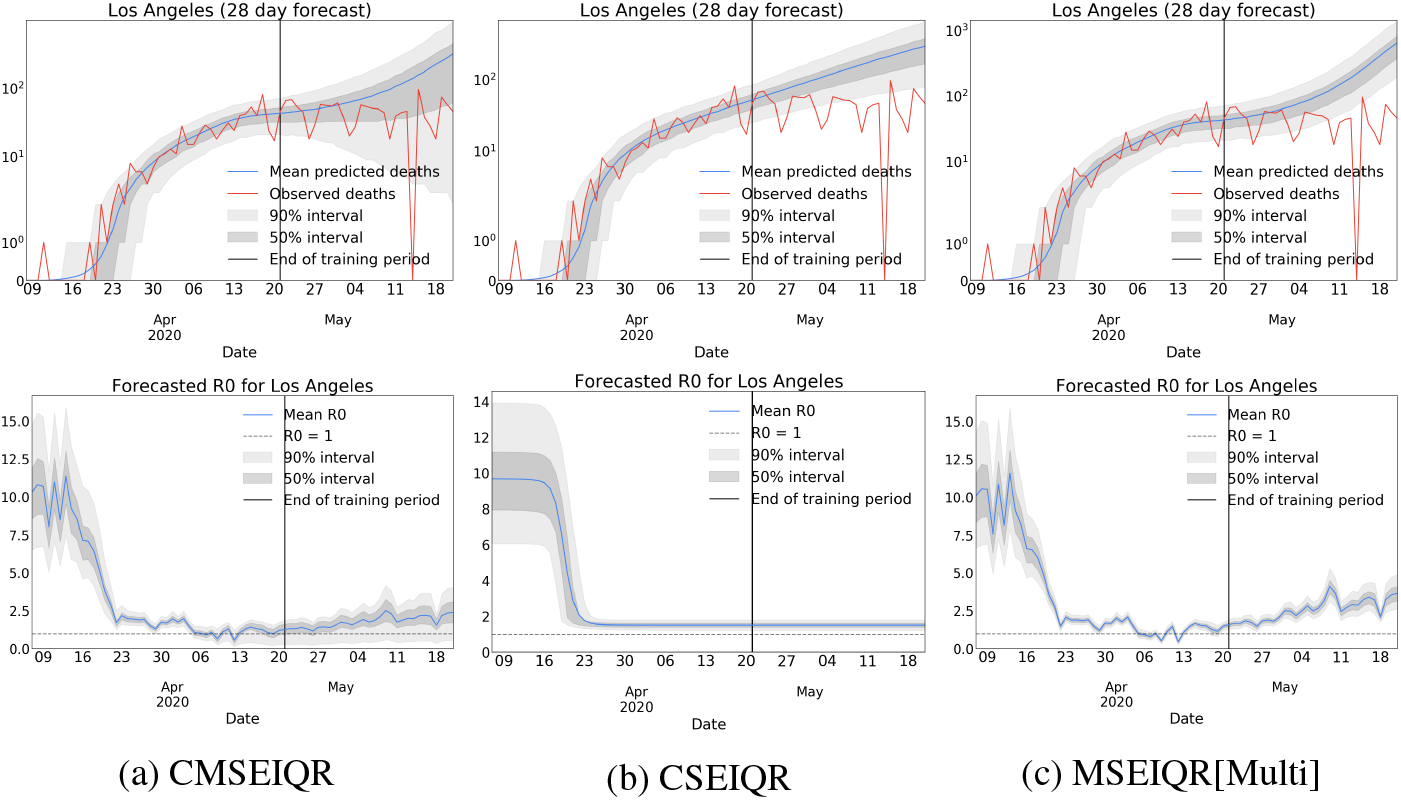
Top row: 28 day death count forecasts for Los Angeles. Bottom row: *R_t_* forecasts for Los Angeles. We observe that the combination of mobility data and a changepoint enables the CMSEIQR model to widen its prediction interval to include a continued flattening of the death curve where other models are unable to predict that pattern.

### 6.2 Nowcasting *R_t_*

Our Bayesian models are able to nowcast the reproductive number *R_t_* for COVID-19 from the posterior and recent mobility data, using the equation: *R_t_* = *β*(*t*) * (1/*η* + *q/γ*), but it is hard to evaluate the accuracy of *R_t_* estimation due to lack of ground truth data. Instead we conduct a held-out-data experiment to validate the consistency and sharpness of a model’s nowcasting results. We pick a date *t*, fit a model trained up to *t*, and compute the distribution over *R_t_*. This model has not seen the deaths that result from infections happening to dates near *t*, so we expect its estimate of *R_t_* to have a wide interval, our “nowcast” estimate. We compare our nowcast distribution to a “hindcast” distribution, a distribution over *R_t_* from a model trained on data up to *t* + 21. In Figure 5 and Figure B.4, we plot a kernel-density estimate of *R_t_* from a range of models. We expect this hindcast distribution has a tighter estimate of *R_t_* since it has access to more data, but we also hope that it lies within the nowcast distribution. We find that CMSEIQR tends to have more conservative but less incorrect nowcasts. After including more data, the CSEIQR and MSEIQR[Multi] models collapse to a sharp estimate, but to an area with low mass in the nowcast, whereas the CMSEIQR model collapses to a less-sharp distribution that is closer to the nowcast. This indicates that the *R_t_* estimates coming from CSEIQR and MSEIQR[Multi] models can be confidently incorrect.

**Figure 5:**
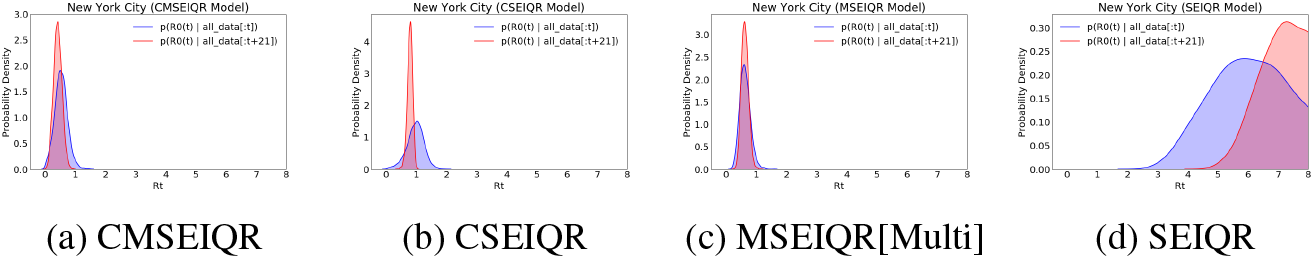
Nowcast vs. hindcast estimates of *R_t_* in New York City on 4/21/20. Nowcast estimates are obtained by training a model on data up to some time *t* and hindcasts are obtained by training a model up to *t* + 21. Nowcasts from the mobility-based models tend to be sharper, but only in CMSEIQR models do we see the nowcast distributions consistent capture the hindcast.

## 7 Discussion

We find that mobility is a promising signal for nowcasting and forecasting the spread of COVID-19, but it is important to understand its limitations. Notably, our experiments suggest that mobility signals alone cannot explain all variation in infection rates; for example, increases in mobility in May do not appear to have caused massive increases in infections. However, we can alleviate these limitations by assuming the existence of and marginalizing other latent factors.

## 8 Broader Impact

It goes without saying that the impact of COVID-19 has been massive and overwhelmingly negative. Our hope is that this paper contributes usefully to the discussion about how best to predict, monitor, and control the spread of COVID-19 using the limited data available. Unfortunately, there is always the danger that work in this space may be misinterpreted or overinterpreted, especially by non-experts. Journalists, policymakers, and politicians may not have the technical training to critically evaluate these models, but they also do not have the luxury of not having an opinion; in the real world, decisions must be made by agents with bounded rationality using imperfect data.

This problem of overinterpretation is particularly salient to this work—it is easy to forget that a model’s error bars are only as reliable as its assumptions. For example, we found that changepoint-only and mobility-only models tend to be overconfident when making long-term forecasts, whereas a more flexible changepoint+mobility model is able to consider a wider variety of scenarios^3^. This should give pause to anyone relying on forecasts from simplistic models for decision-making— “simpler” does not necessarily imply “fewer assumptions” or “more reliable”.

All of this is cause for caution, but not inaction. Even imperfect models are still useful tools for rigorously and honestly integrating our prior beliefs and the limited evidence we can collect, as long as we remember that they *are* imperfect.

## Data Availability

We use unprocessed COVID-19 case and death count numbers from the New York Times, which produces daily reports of new infections and deaths at both state and county level. For mobility, we use the Community Mobility Reports published by Google in our evaluation of Bayesian compartmental models.

https://www.nytimes.com/interactive/2020/us/coronavirus-us-cases.html

https://www.google.com/covid19/mobility/

## A Prior distributions

We report our choice of prior distributions in Table A.2.

**Table A.2:** Prior distributions used when defining Bayesian models. Priors were informed by surrounding literature and in some cases were constrained (i.e. the use of LogitNormal) to avoid the inference machinery finding degenerate parameter settings that exploit numerical issues in the simulator.

| Parameter | Prior | Notes |
| --- | --- | --- |
| $\beta$ (base infection rate) | LogNormal(0, 0.5) | |
| $\alpha$ (incubation rate) | LogitNormal(logit(1/5.), .1) | |
| $\eta$ (isolation rate) | LogitNormal(logit(1/3.), 2.) | |
| $\gamma$ (recovery rate) | LogNormal($-\log 16$, 0.1) | |
| $q$ (quarantine reduction) | Uniform(0, 0.1) | |
| $w_c$ (city mobility coefficient) | LogNormal(0.5, 1) | Mobility models only |
| $f$ (mobility feature adjustments) | $\mathcal{N}(0, 1)$ | Mobility models only |
| $b$ (mobility intercept) | $\mathcal{N}(0, I)$ | Mobility models only |
| $\kappa$ (changepoint time) | Uniform(0, 1) | Changepoint models only |
| $\rho$ (changepoint magnitude) | Uniform(0, 1) | Changepoint models only |
| $\nu$ (changepoint rate) | LogNormal(1, 1) | Changepoint models only |
| $\omega$ (infection fatality ratio) | LogitNormal(logit(0.007), 0.1) | Likelihood parameter |
| $r$ (neg-binomial shape parameter) | LogNormal(log 30, 0.1) | Likelihood parameter |

## B SMC results

### B.1 Trace plots

**Figure B.2:**
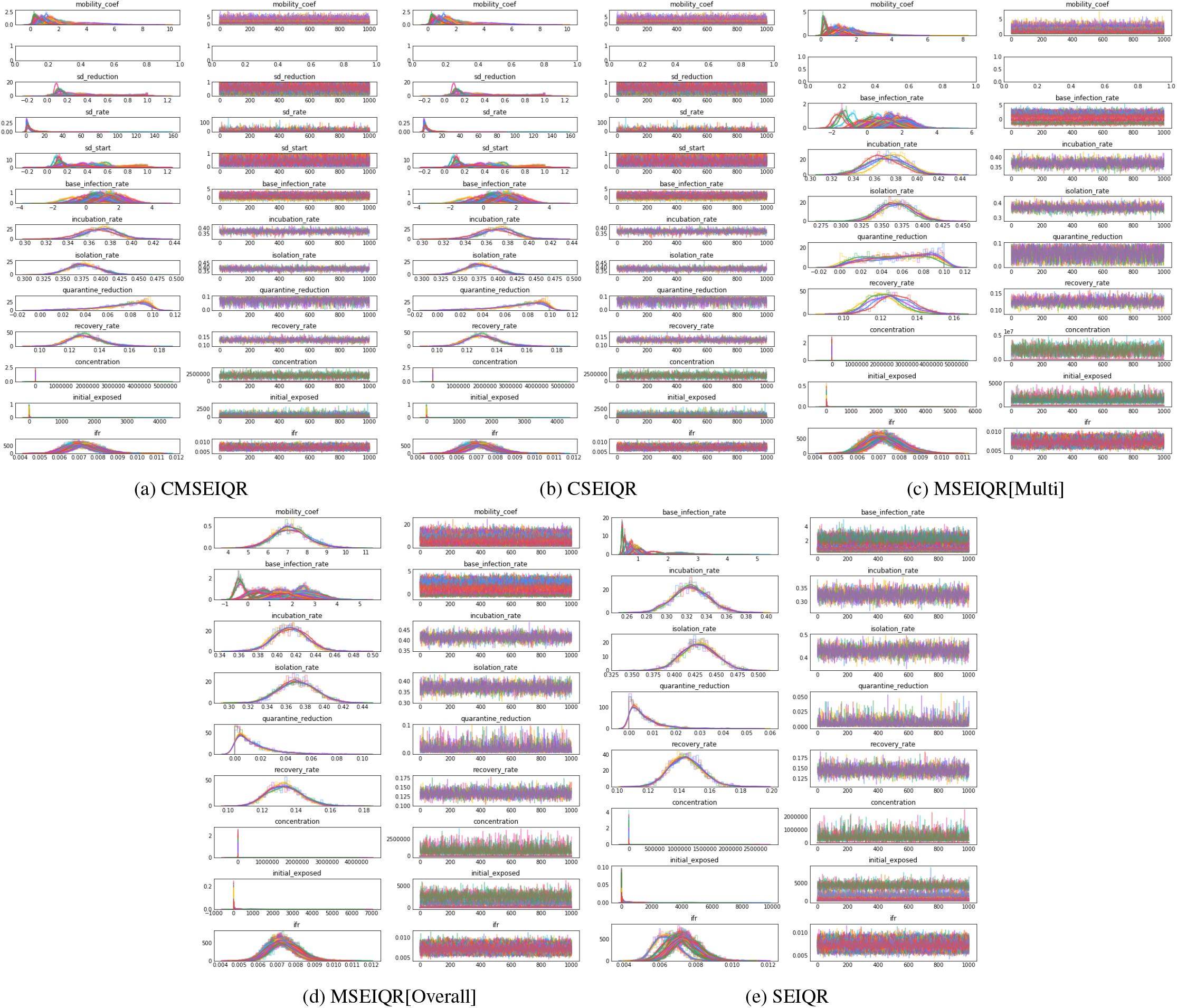
Trace plots for SMC inference.

**Figure B.3:**
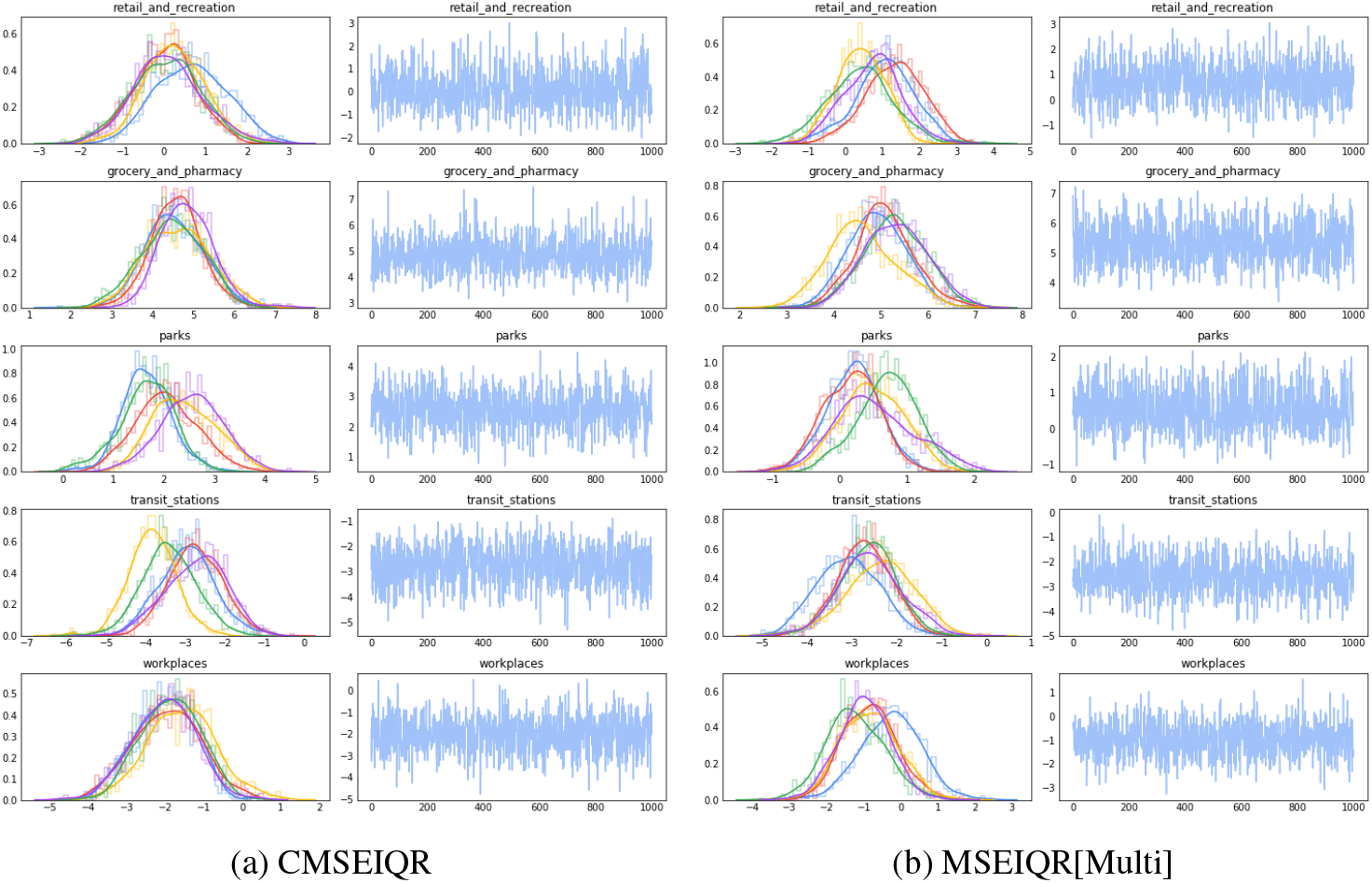
Trace plots for the posterior over mobility feature adjustments.

**Figure B.4:**
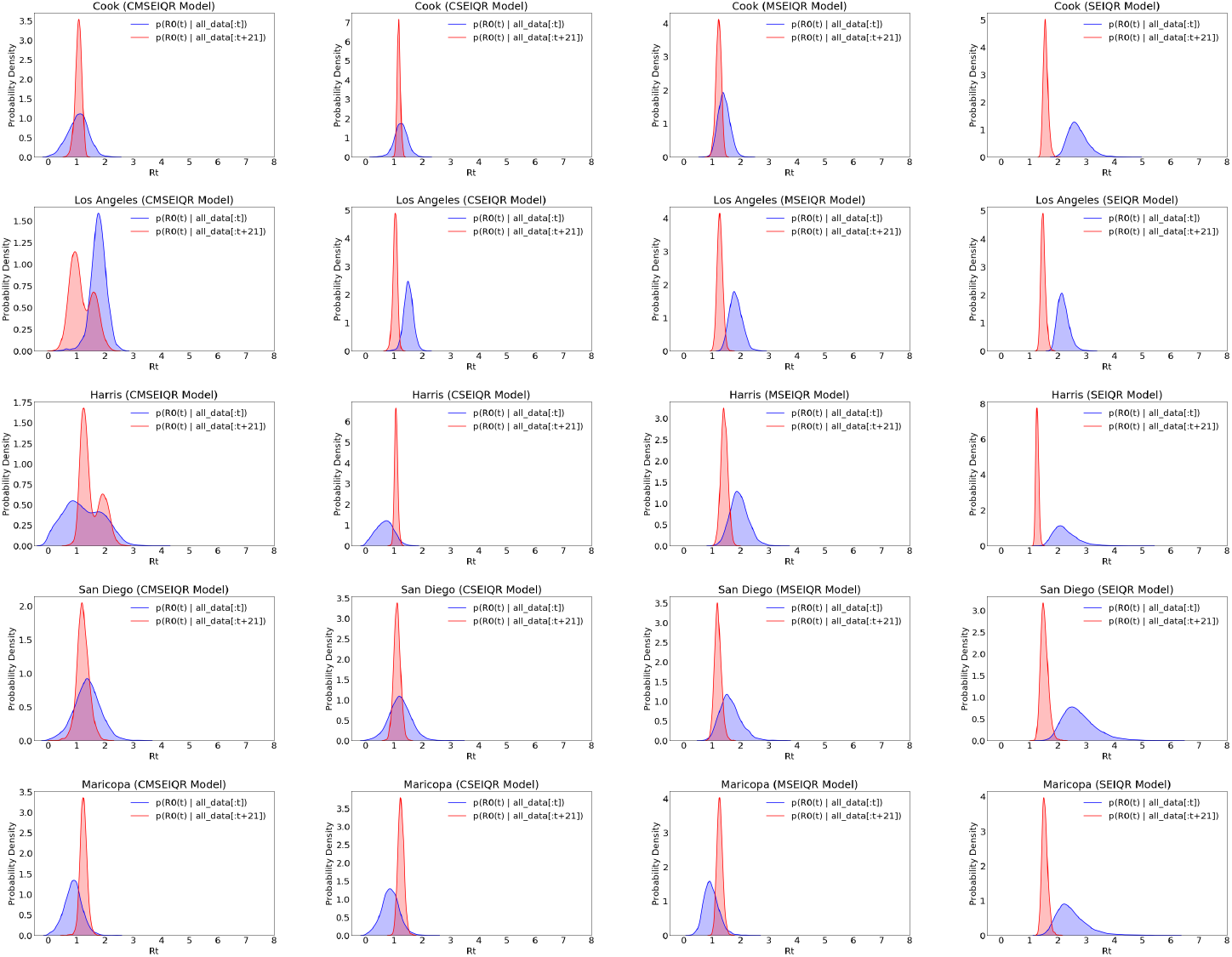
Nowcasted versus hindcasted *R_t_* in 5 counties on 4/21/20.

### B.2 Average log-likelihood tables

**Table B.3:**
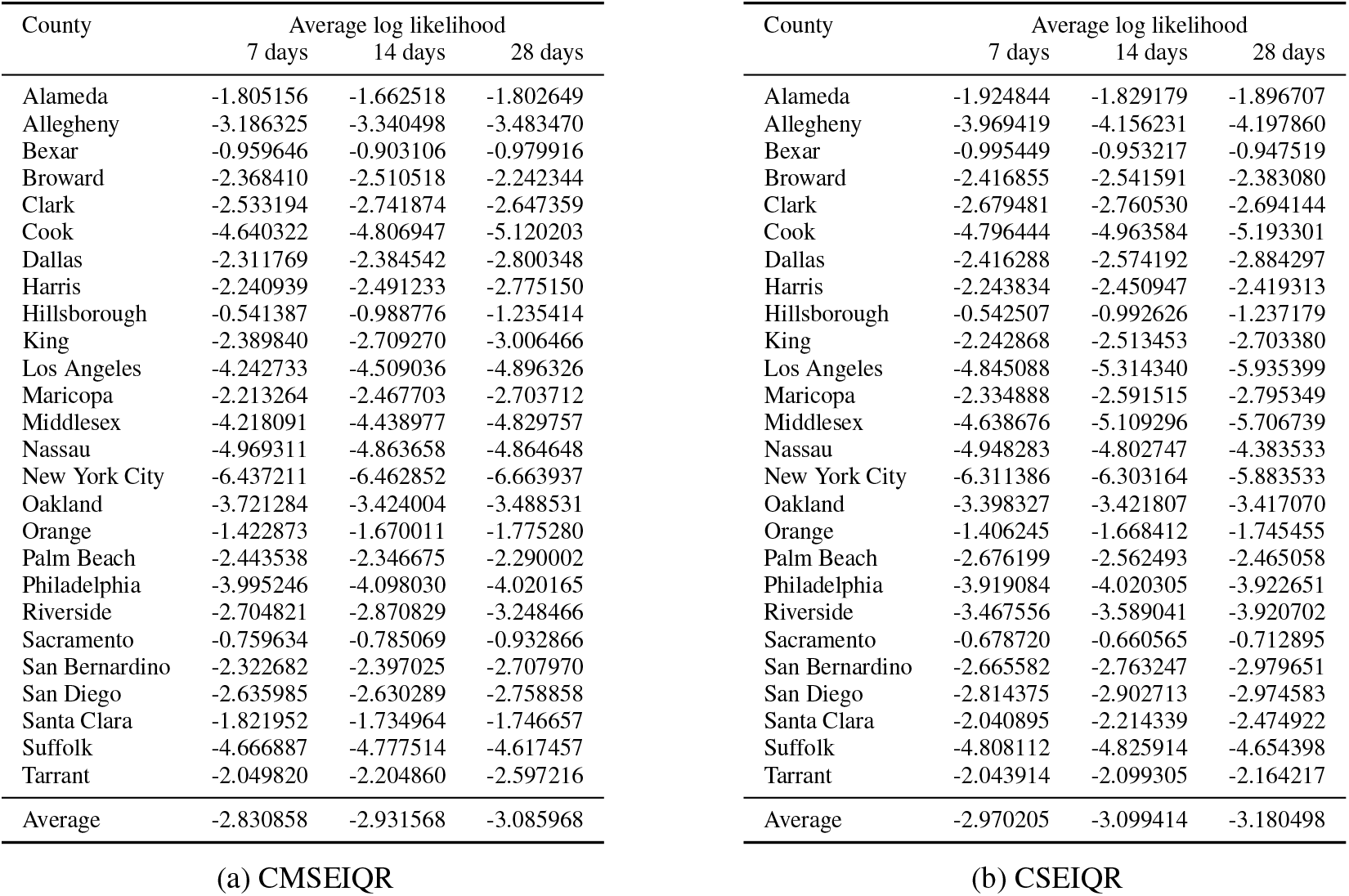
Average held-out log-likelihood numbers for the CMSEIQR and CSEIQR models.

**Table B.4:**
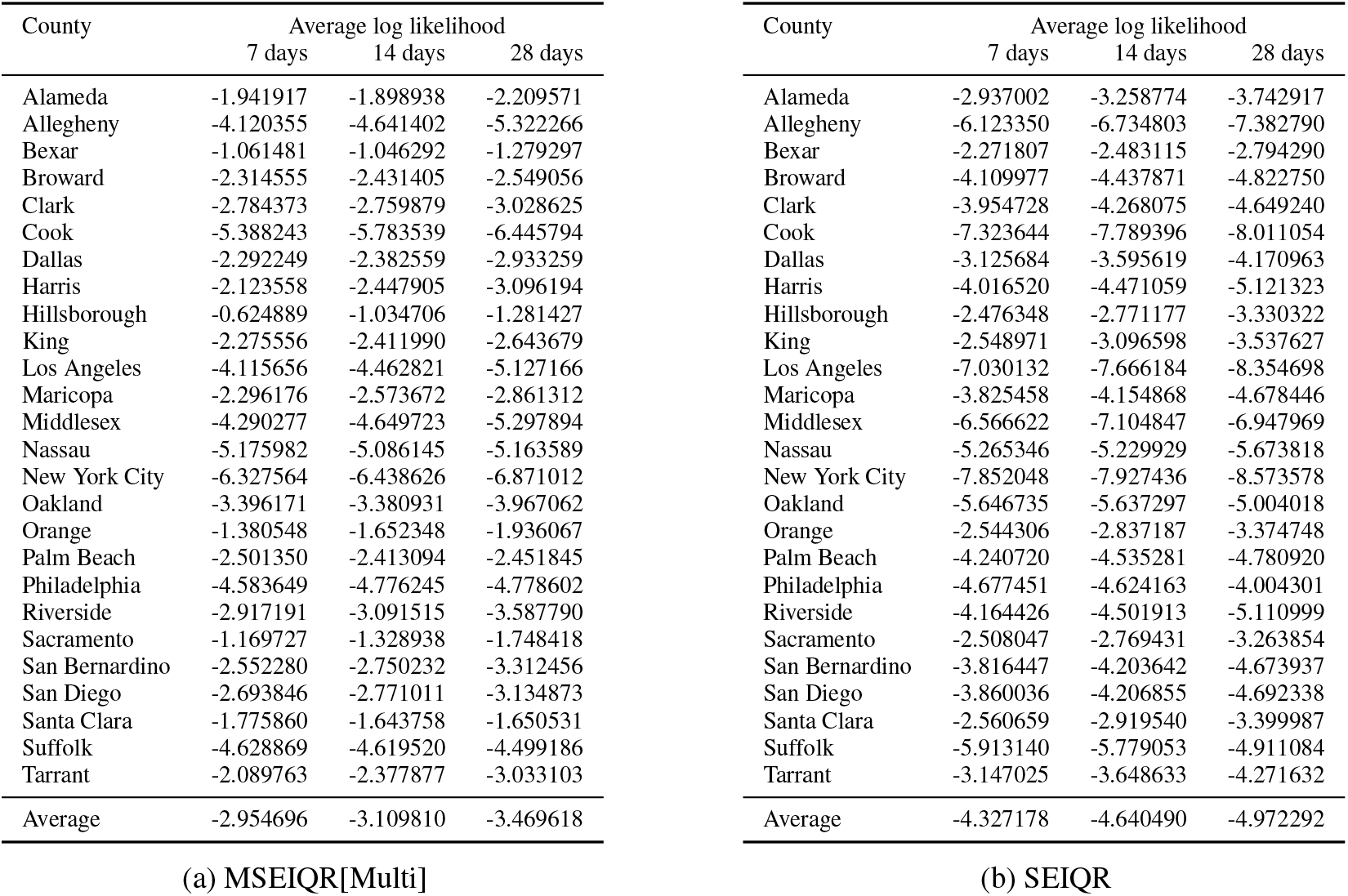
Average held-out log-likelihood numbers for the MSEIQR[Multi] and SEIQR models.

**Table B.5:** Average held-out log-likelihood numbers for MSEIQR[Overall].

| County | Average log likelihood |  |  |
| --- | --- | --- | --- |
|  | 7 days | 14 days | 28 days |
| Alameda | -2.272243 | -2.472632 | -2.921608 |
| Allegheny | -5.290524 | -5.886054 | -6.561562 |
| Bexar | -1.243805 | -1.303534 | -1.624272 |
| Broward | -2.475610 | -2.601691 | -2.922158 |
| Clark | -2.874675 | -2.874134 | -3.124725 |
| Cook | -5.792146 | -6.284522 | -6.955929 |
| Dallas | -2.441165 | -2.762431 | -3.360741 |
| Harris | -2.133428 | -2.422409 | -3.047185 |
| Hillsborough | -0.825620 | -1.166549 | -1.419374 |
| King | -2.240625 | -2.512409 | -2.757119 |
| Los Angeles | -4.267687 | -4.723704 | -5.405117 |
| Maricopa | -2.463335 | -2.714478 | -2.977257 |
| Middlesex | -4.919390 | -5.490939 | -6.170833 |
| Nassau | -5.413037 | -5.358217 | -5.134870 |
| New York City | -6.433594 | -6.645010 | -7.161345 |
| Oakland | -3.410262 | -3.364388 | -3.953322 |
| Orange | -1.462976 | -1.734681 | -2.040418 |
| Palm Beach | -2.711717 | -2.611960 | -2.749688 |
| Philadelphia | -4.698258 | -4.810053 | -4.510210 |
| Riverside | -3.496074 | -3.653838 | -4.152404 |
| Sacramento | -1.288776 | -1.481599 | -1.875778 |
| San Bernardino | -2.881672 | -3.168224 | -3.732581 |
| San Diego | -2.868105 | -3.010828 | -3.412882 |
| Santa Clara | -2.058977 | -2.258750 | -2.665601 |
| Suffolk | -4.725725 | -4.737740 | -4.986308 |
| Tarrant | -2.140717 | -2.446194 | -3.086203 |
| Average | -3.185775 | -3.403730 | -3.796519 |

## C Additional plots

### C.1 Additional forecasts

**Figure C.5:**
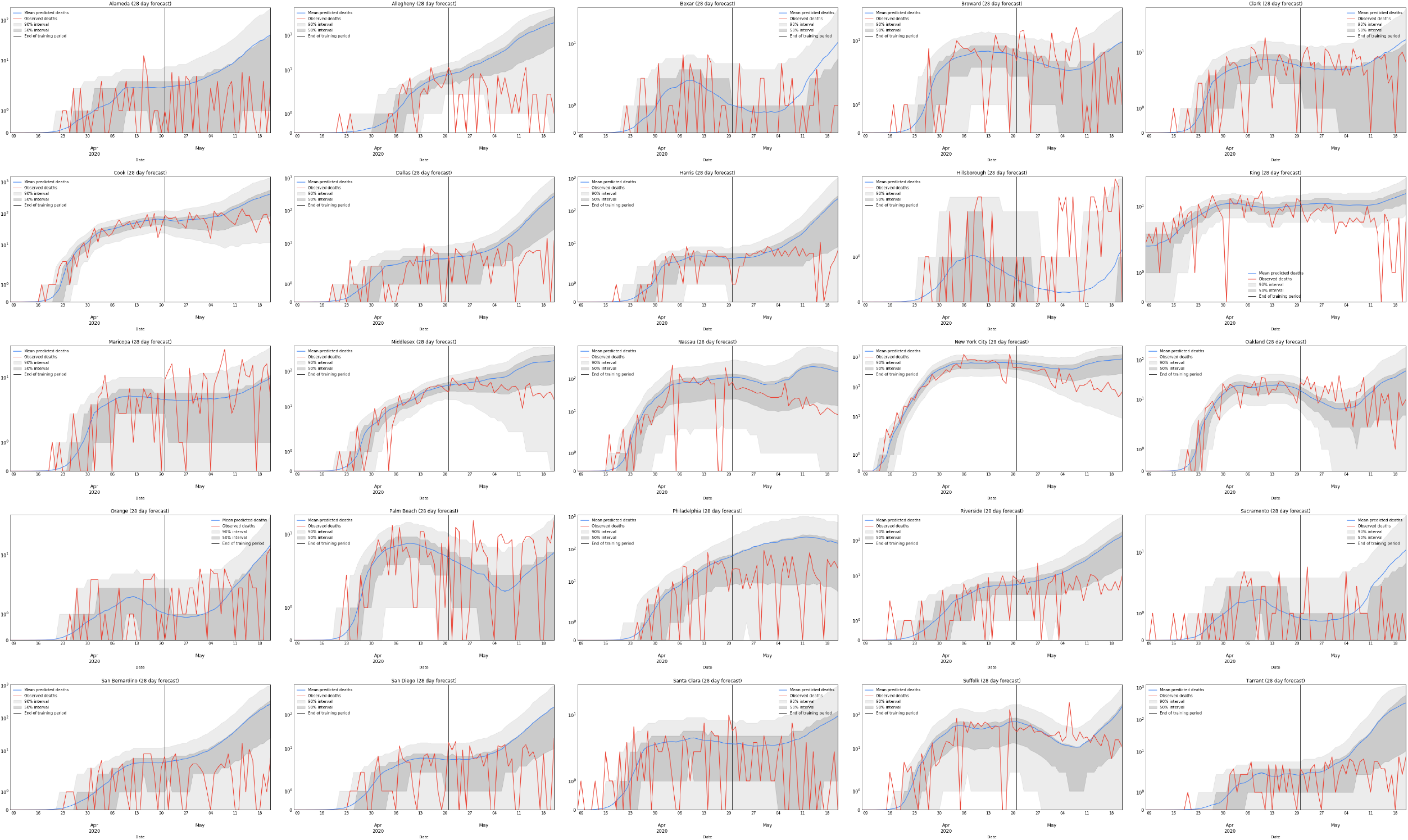
All forecasts for CMSEIQR except Los Angeles.

**Figure C.6:**
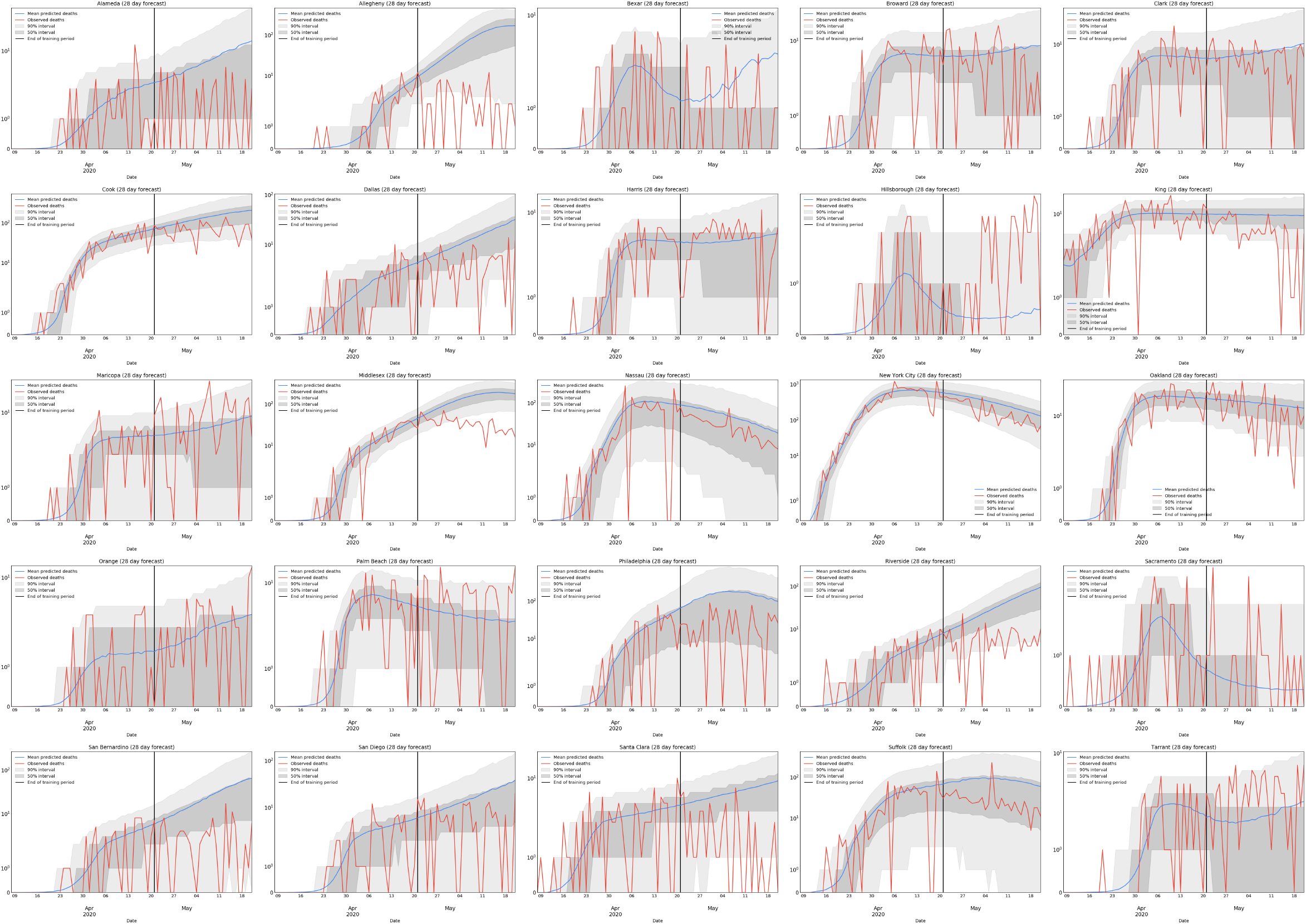
All forecasts for CSEIQR except Los Angeles.

**Figure C.7:**
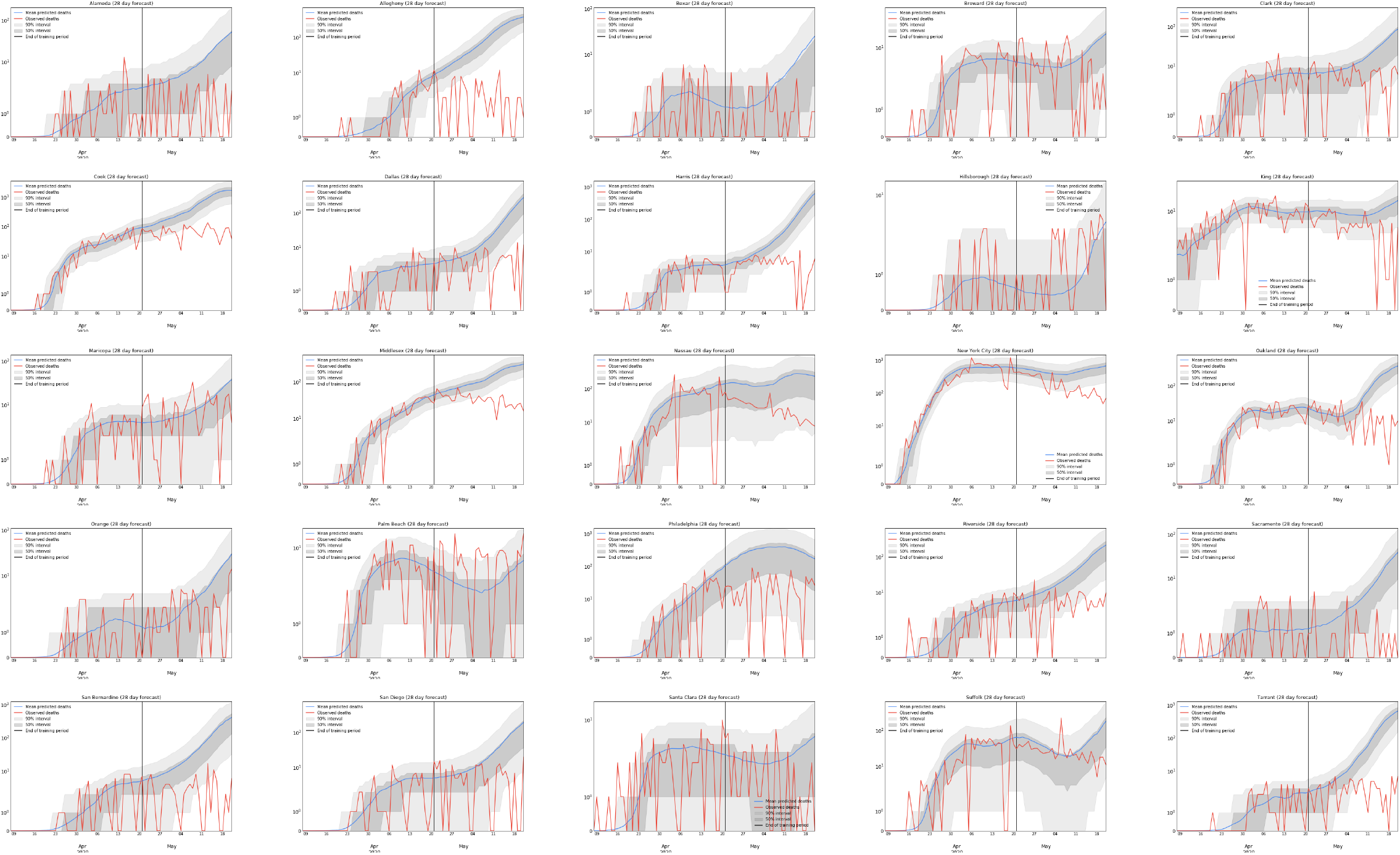
All forecasts for MSEIQR[Multi] except Los Angeles.

**Figure C.8:**
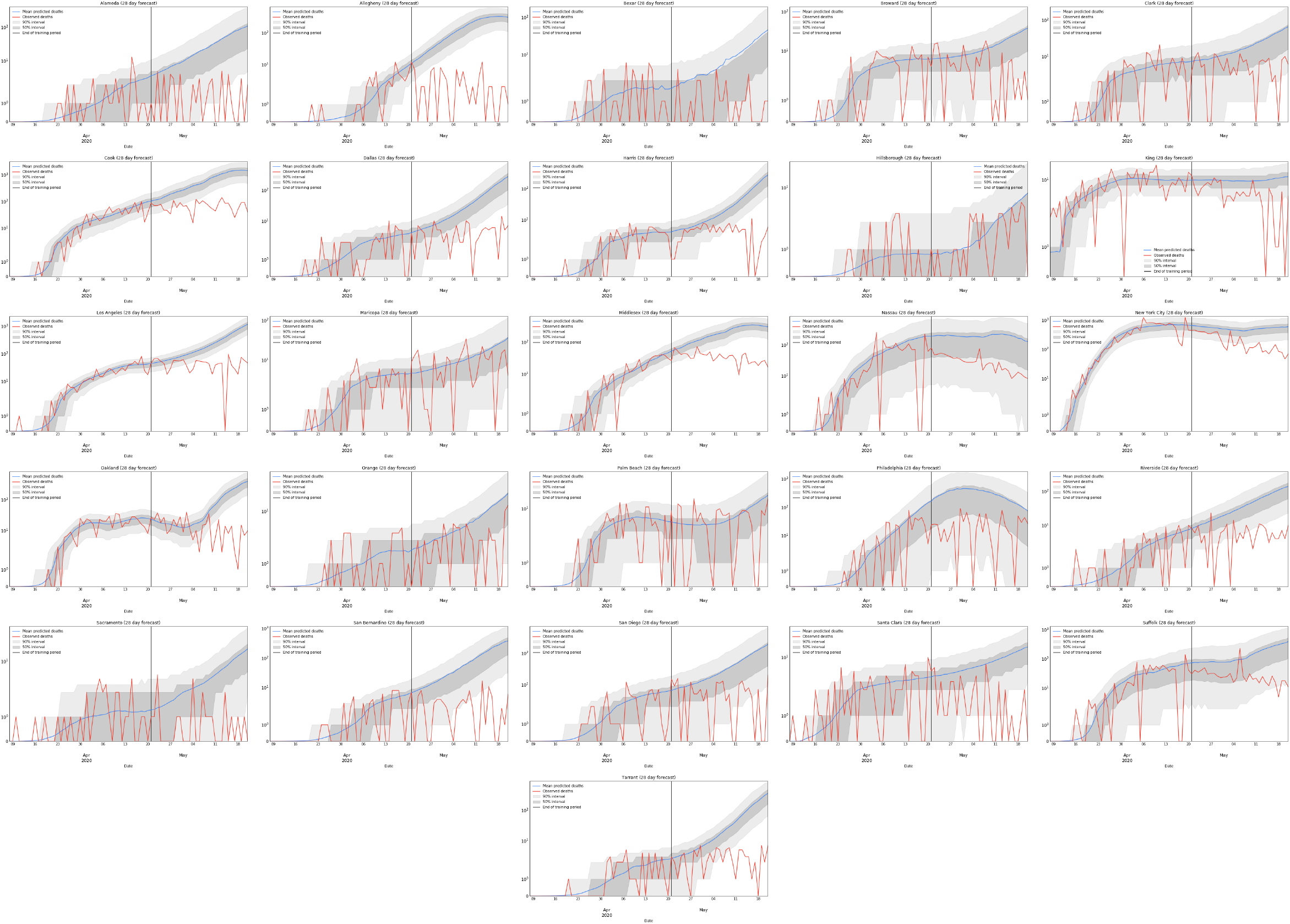
All forecasts for MSEIQR[Overall].

**Figure C.9:**
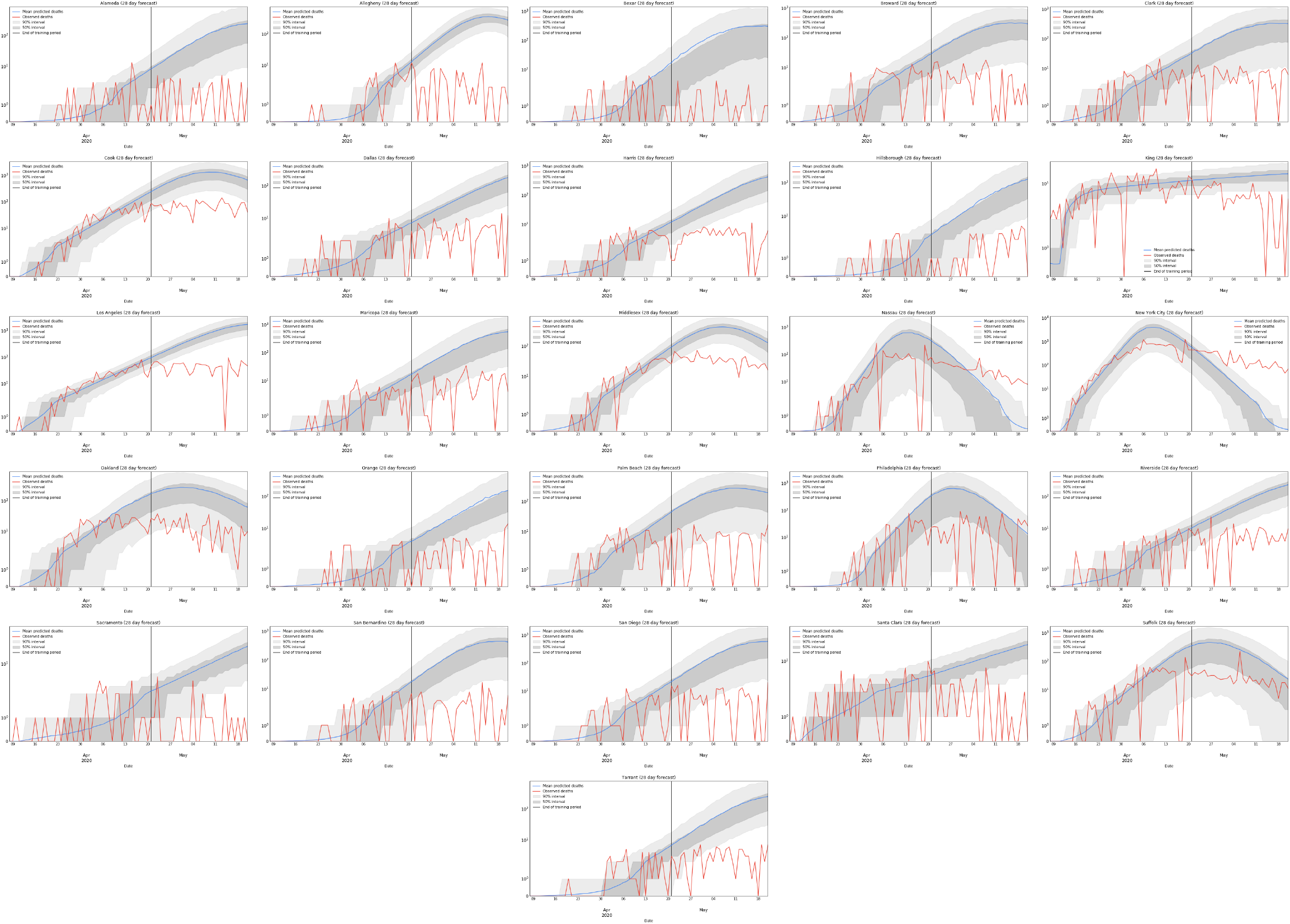
All forecasts for SEIQR.

### C.2 Additional *R_t_* forecasts

**Figure C.10:**
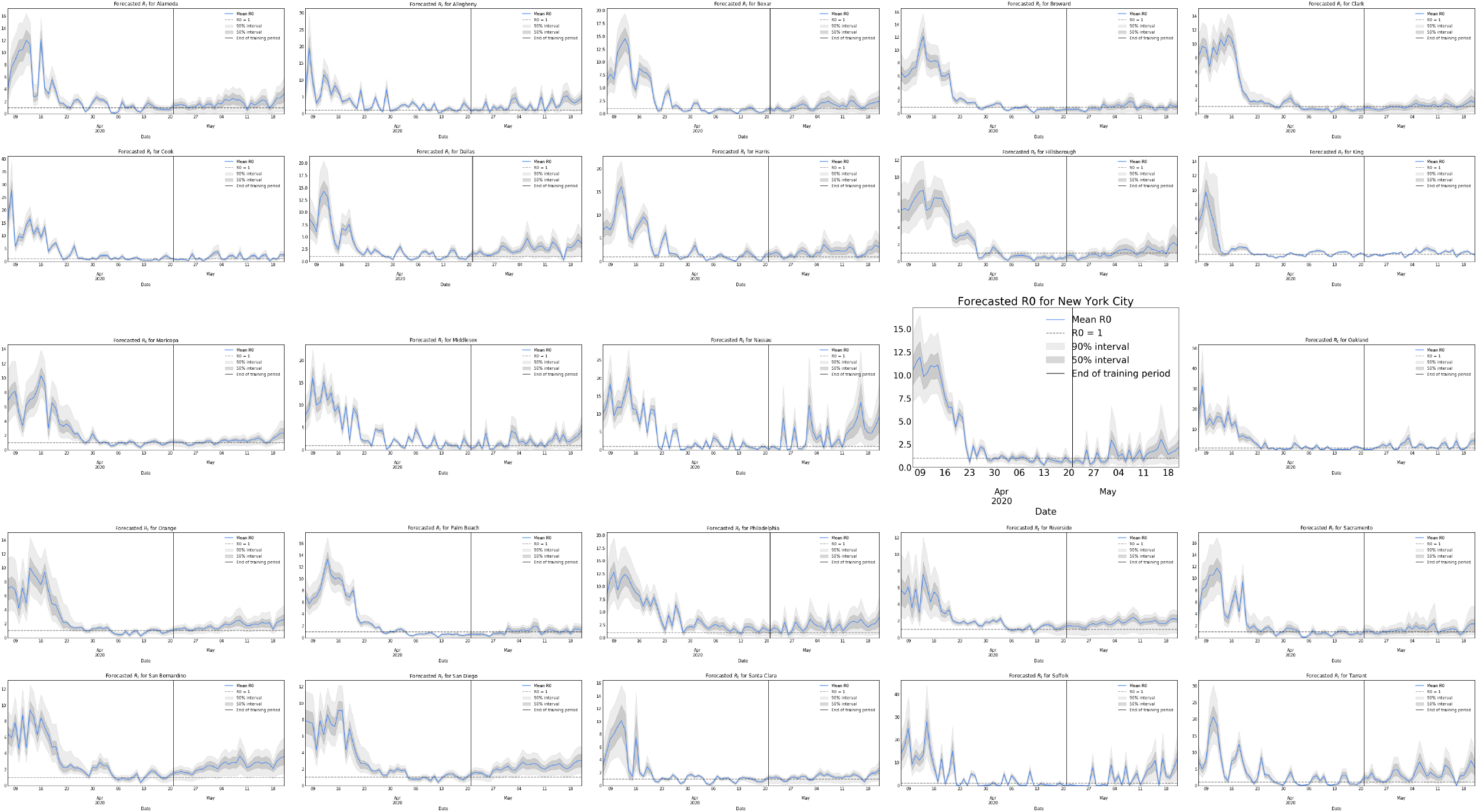
All *R_t_* forecasts for CMSEIQR except Los Angeles.

**Figure C.11:**
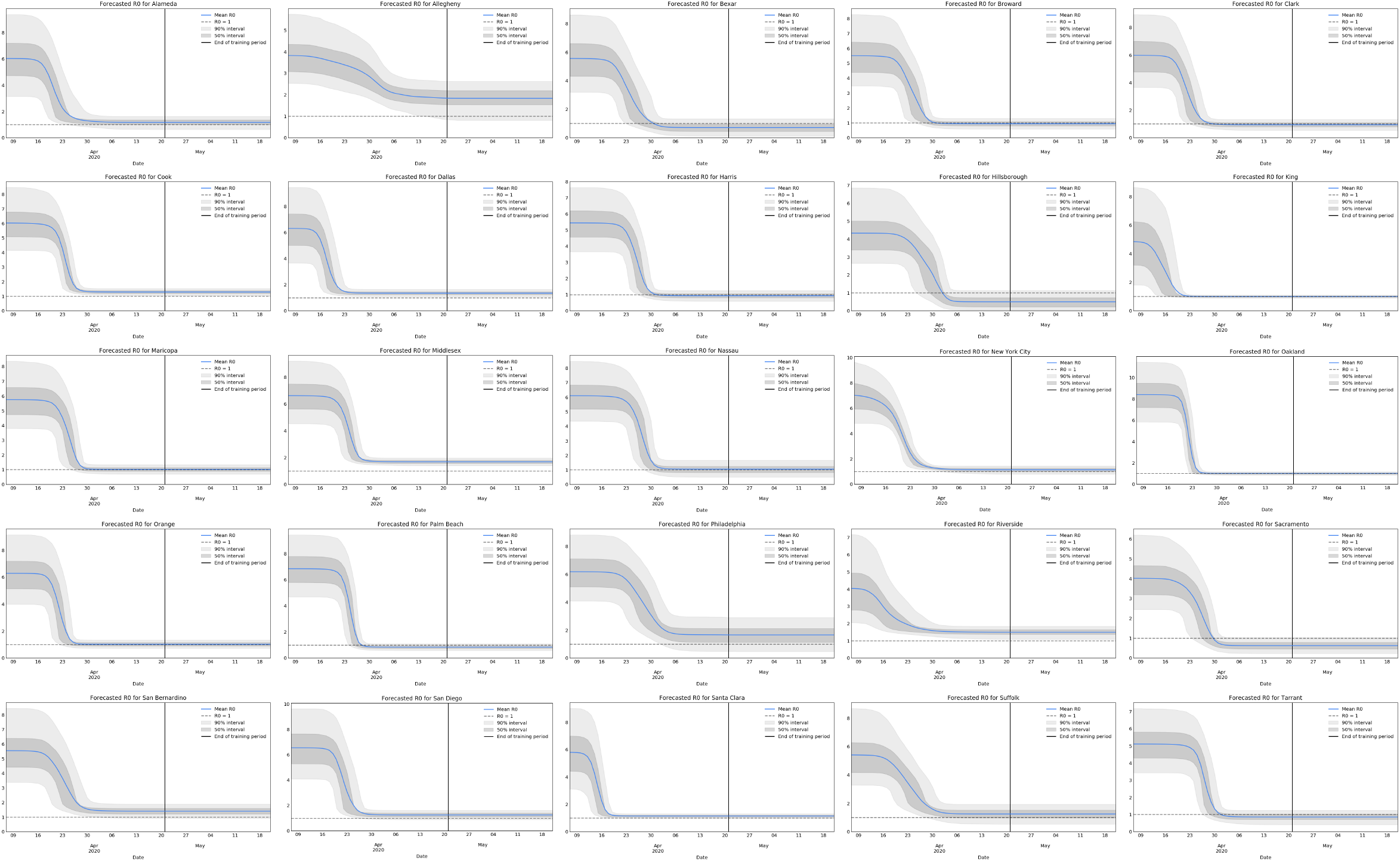
All *R_t_* forecasts for CSEIQR except Los Angeles.

**Figure C.12:**
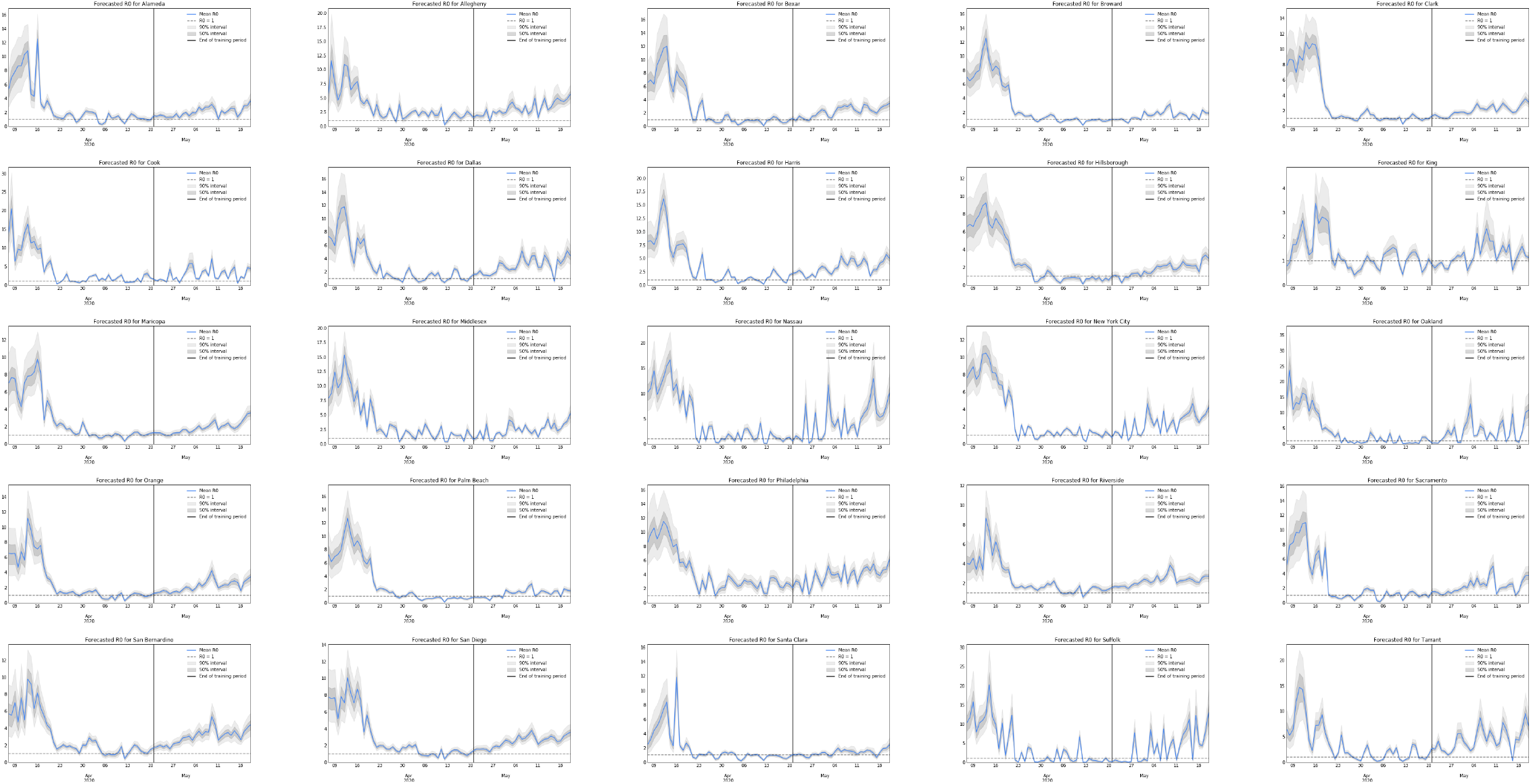
All *R_t_* forecasts for MSEIQR[Multi] except Los Angeles.

**Figure C.13:**
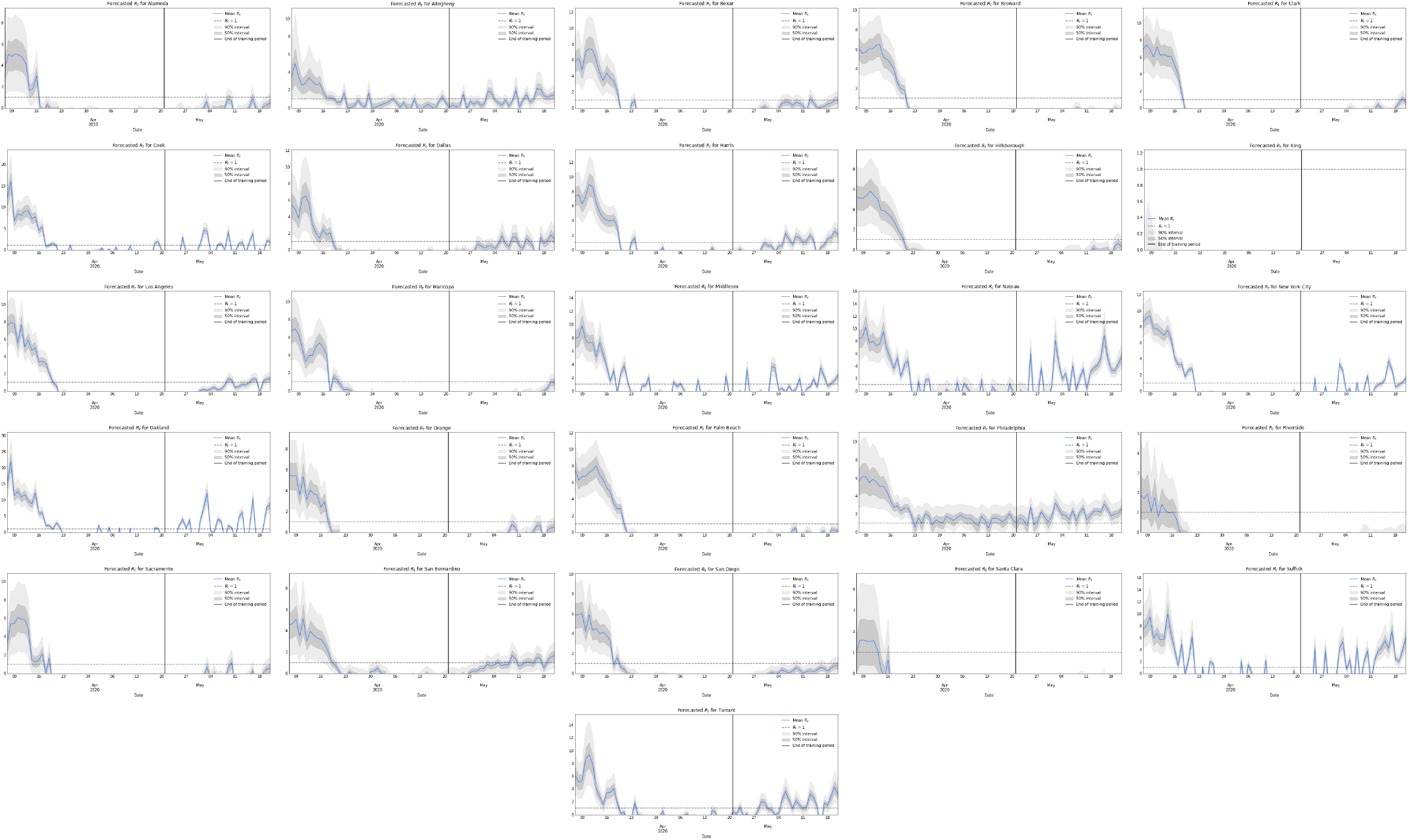
All *R_t_* forecasts for MSEIQR[Overall].

**Figure C.14:**
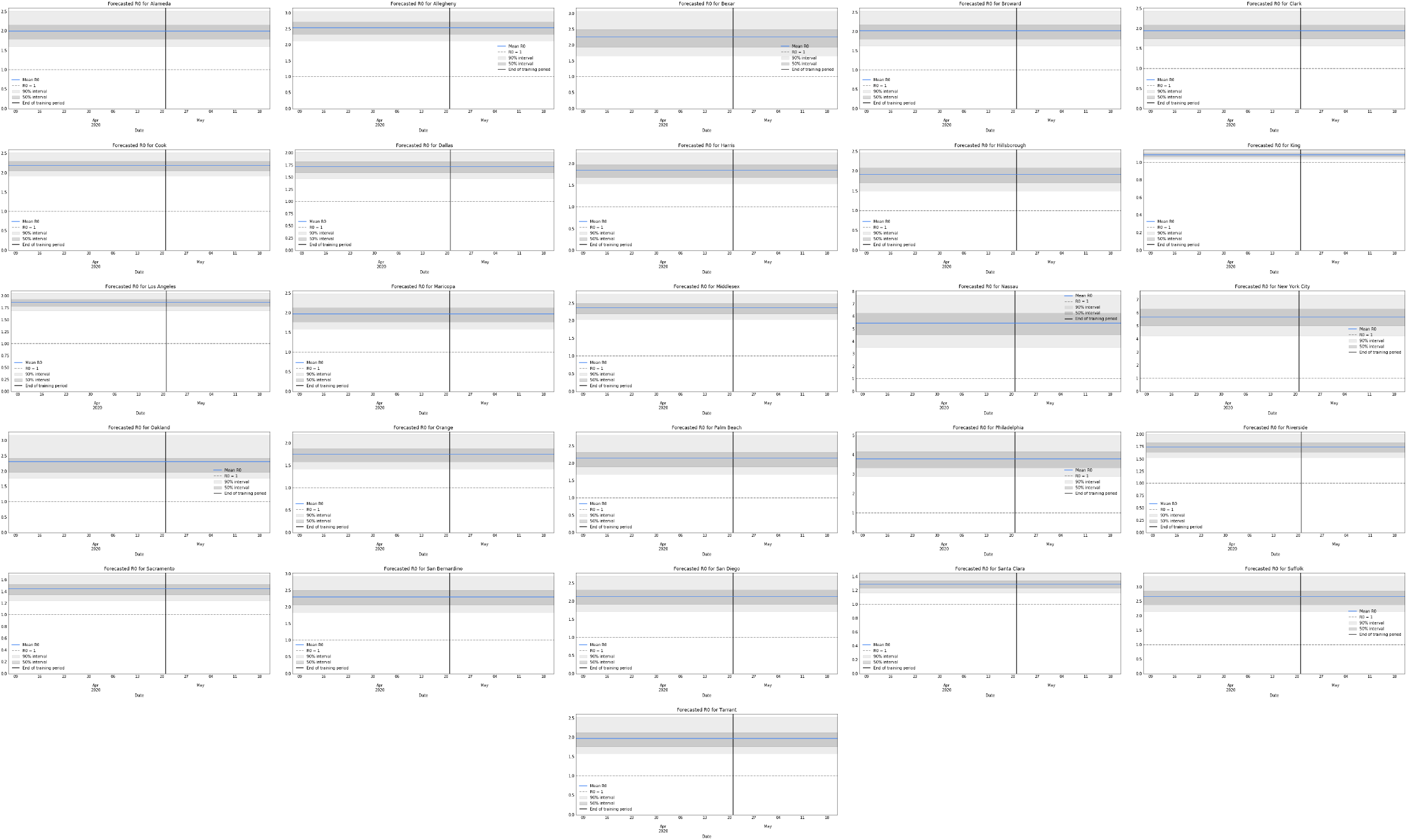
All *R_t_* forecasts for SEIQR.

## Footnotes

2 In a real forecasting application, one would also need a forecast of how mobility would change over the forecast period; this experiment assumes a perfect mobility forecast. Note that the influence of this mobility forecast should be relatively minor due to the weeks-long lag between infections and deaths.

3 Note that this example ignores the difficulty of predicting (let alone controlling) future mobility trends and behavior.

